# Multiomic profiling of metastatic potential in estrogen receptor-positive human epidermal growth factor-negative breast cancer

**DOI:** 10.1101/2025.01.22.25320944

**Authors:** Sergio Mosquim, Måns Zamore, Johan Vallon-Christersson, Lisa Rydén, Fredrik Levander

**Affiliations:** Department of Immunotechnology, Lund University, Sweden; Division of Oncology, Department of Clinical Sciences Lund, Lund University; Division of Surgery, Department of Clinical Sciences Lund, Lund University, Lund; Department of Surgery, Skåne University Hospital, Lund, Sweden; National Bioinformatics Infrastructure Sweden (NBIS), Science for Life Laboratory, Lund University, Sweden

## Abstract

Metastatic disease is the main cause of breast cancer-related deaths. Given advances in the molecular profiling of tumors, we here aimed at integrating proteome, phosphoproteome and transcriptome data for the profiling of primary tumors to allow for discovery of new subtypes and features predicting lymph node and distant metastases in breast cancer (BC). We analyzed a total of 182 estrogen-receptor (ER) positive, human epidermal growth factor receptor 2 (HER2) negative BC samples using label-free Data Independent Acquisition (DIA) liquid chromatography tandem mass spectrometry (LC-MS/MS), quantifying a total of 13571 protein groups, 7107 phosphopeptides and 13085 expressed genes in at least 70% of the samples. Unsupervised consensus cluster analyses permitted the identification of potential subtypes with differential immune infiltration pattern and survival. Immune deconvolution data was combined with multiomics factor analysis, providing unique insight into different markers for lymph node and distant metastases. In summary, we further developed a protocol for parallel acquisition of matched proteomics, phosphoproteomics and transcriptomics data, resulting in the most comprehensive dataset of its kind and allowing for unique insights into metastatic processes in ER-positive/HER2-negative BC.

## Introduction

According to statistics from the Global Cancer Observatory, in 2022, Breast Cancer (BC) was responsible for nearly one quarter of all cancer diagnoses in women. It also ranks first in cancer mortality for women, with 666 103 deaths registered in 2022^1–5^. Continuous advances both in terms of genomic and transcriptomic characterization, as well as early detection by public screening programs, have contributed to a reduction in the mortality rate and improved patient stratification^6–8^.

Metastasis, one of the hallmarks of cancer described by Hanahan^9^ is the main cause of death amongst cancer patients ^9–12^. The metastatic dissemination is a process which can take place via blood vessels and lymphatic vessels^11^. In BC, the lymphatic system is thought to be crucial for metastatic spread, where the presence of lymph node metastases is not only the earliest sign of metastatic spread, but it also constitutes an important prognostic marker^10,13,14^.

In BC, lymph node staging is a crucial part of the diagnostic process^14^. Clinically, the initial assessment of axillary lymph node (ALN) status is commonly performed via ultrasound. However, since imaging alone is not enough to rule out negative cases, these patients are further submitted to sentinel lymph node (SLN) biopsy, recommended for clinically node-negative cases^14–16^. If the results of SLN biopsy (SLNB) are positive, then completion axillary node dissection (cALND) would previously be performed^14^.

Originally, cALND was designed for staging and improving the overall survival of patients. However, the procedure is also associated with several side effects. The proposal of SLNB came as an alternative to minimize such side effects, supported by the results from different clinical trials which demonstrated that SLNB was not inferior to cALND in terms of survival^14,15,17^. In recent years, however, the use of SLNB has been questioned. Although SLNB is the standard procedure for ALN evaluation in node negative disease, it is still associated with reduced quality of life, besides having negative findings in 70% to 80% of cases and a well-established false negative rate of 10%, rendering the procedure mostly diagnostic and not therapeutic^5,14–16^.

Molecular profiling of tumor biopsies is becoming standard practice and is guiding treatment along with clinical parameters. Several biomarkers are used for cancer classification, and it is feasible to believe that protein activity profiles could be used also to predict tumor spread. Transcriptional profiling has indeed added predictive power to lymph node prediction, although still at a limited scale. With protocols enabling simultaneous proteomics and transcriptomics profiling of tumors, multiomics profiling could thus further improve our understanding of metastatic potential through lymph node involvement, with possible clinical implications.

Here, we build on a previously developed protocol for characterization of tumors through transcriptomics and proteomics and include phosphoproteomics to study a selection of samples from the SCAN-B cohort^18^. The aim is to detect molecular characteristics of tumors with metastatic potential, focusing on the largest group of breast cancer patients, i.e., estrogen-receptor (ER) positive and human epidermal growth factor receptor 2 (HER2) negative tumors, with a relatively good prognosis.

## Results

### Acquisition of multiomics data from breast cancer tissue

In the present study, we aimed at investigating a multiomic profile of metastatic processes in estrogen receptor positive BC. A high throughput protocol allowing for parallel RNA and protein measure was previously described ^19^. Here, we further developed the protocol to include parallel phosphoproteome data acquisition.

A total of 182 BC samples belonging to the SCAN-B cohort^18^ were used in the present study. Based on immunohistochemistry (IHC) results, the samples were selected to be ER-positive HER2-negative. As the two main histological classifications of invasive BC are invasive breast carcinoma of no special type (IBC-NST) – or just non-special type (NST) carcinomas – and invasive lobular (ILC), samples of both subtypes were selected. A graphical representation of the study design can be seen in **Figure 1**.

**Figure 1.**
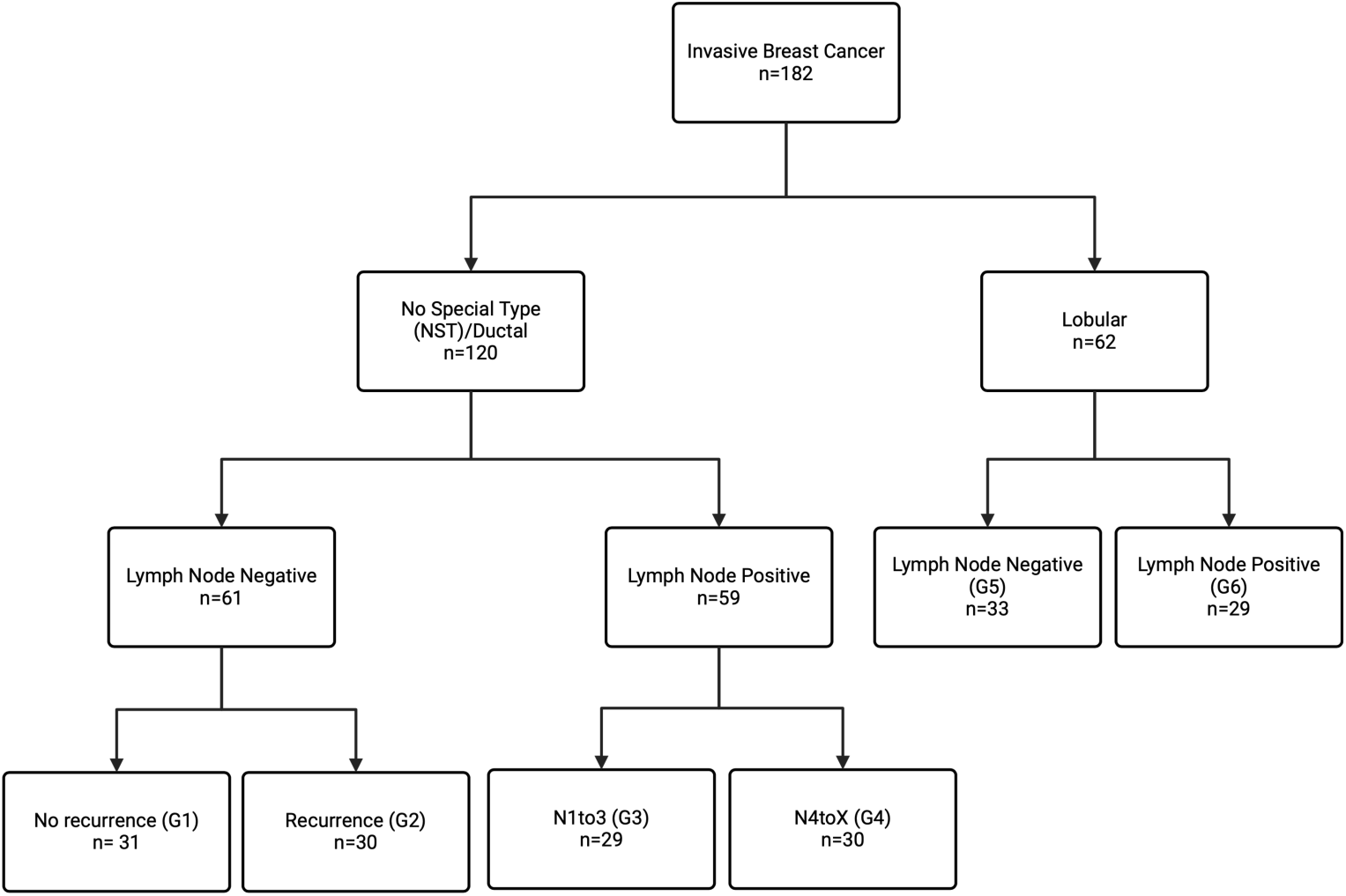
Flowchart representing the study design. A total of 182 breast cancer samples were selected, 120 of which are NST and the remaining 62 are ILC. The samples were further stratified according to lymph node status. Due to the higher incidence of NST, the lymph node subgroups were further stratified according to the absence or presence of distant recurrence event (Group1 and Group2, respectively), and according to nodal burden, 1to3 in Group3 and 4toX in Group4. Groups 5 and 6 represent lymph node negative and positive ILC cases, respectively. Created with BioRender.com.

The input sample material corresponds to the flowthrough after DNA and RNA extraction via the implementation of the AllPrep protocol in the Qiagen cube, described in ^18^. These flowthroughs contain the protein fraction, except for proteins potentially lost during previous extraction steps. As the input material comes from the same original biopsy collected, our current protocol allows for matched transcriptome, proteome and phosphoproteome data. To the best of our knowledge, this is the most comprehensive matched proteomic and phosphoproteomic dataset of such nature, allowing for a unique insight into the metastatic processes in ER-positive HER2-negative BC.

The Data Independent Acquisition (DIA) strategy used allowed for identification of a total of 17860 protein groups and 26150 phosphopeptides (**Table 1**). These data were further combined with previously acquired transcriptomics data ^20^ and studied using integrative omics methods. To validate the potential of the approach, we first conducted characterization of histological subtypes with known molecular differences, before exploring molecular differences of lymph node involvement and distant metastasis.

**Table 1:** Overview of the number of features identified in all three omics layers.

| OMICS | Number of Features | At Least 70% D.C. | Complete observations |
| --- | --- | --- | --- |
| Proteomics DIA (protein groups) | 17860 | 13571 | 4426 |
| Phosphoproteomics DIA (peptides) | 26150 | 7107 | 1329 |
| Transcriptomics (genes) | 19675 | 13085 | 13065 |

D.C.: Data Completeness

### Key differences between invasive ductal and lobular carcinomas

Due to the heterogeneity of BC, different histological subtypes have been described^21^. However, the two most common are NST and ILC^21–23^. These subtypes differ at the clinical, histological and molecular level^23^.

The ILC subtype corresponds to around 10-15% of all BC cases^21,22^, and in our selected cohort ^20^, further filtered to focus on ER-positive HER2-negative cases, ILC represents approximately 17% of cases. ILC differs from NST in that cases are normally less proliferative, of lower grade, and the vast majority, over 90%, presents as ER-positive and HER2-negative^21–23^. Morphologically, ILC is characterized by having small discohesive cells that grow in a single file. As a result, cases are less likely to form masses, which makes them more difficult to be detected via conventional imaging techniques ^21–23^.

The hallmark of ILC, and what drives these morphological differences, is the loss of E-Cadherin (CDH1) expression^22^. This loss is observed in about 90% of ILCs and can be used in the context of immunohistochemistry to aid in the classification of borderline cases, i.e., those that have mixed histological features^22^.

As a way of validating the data collected in the present study, we investigated whether differentially abundant features between NST and ILC were in line with the biological characteristics of these subtypes. To achieve that, we performed differential expression analysis between NST and ILC at the proteome, phosphoproteome and transcriptome levels (**Figure 2**).

**Figure 2.**
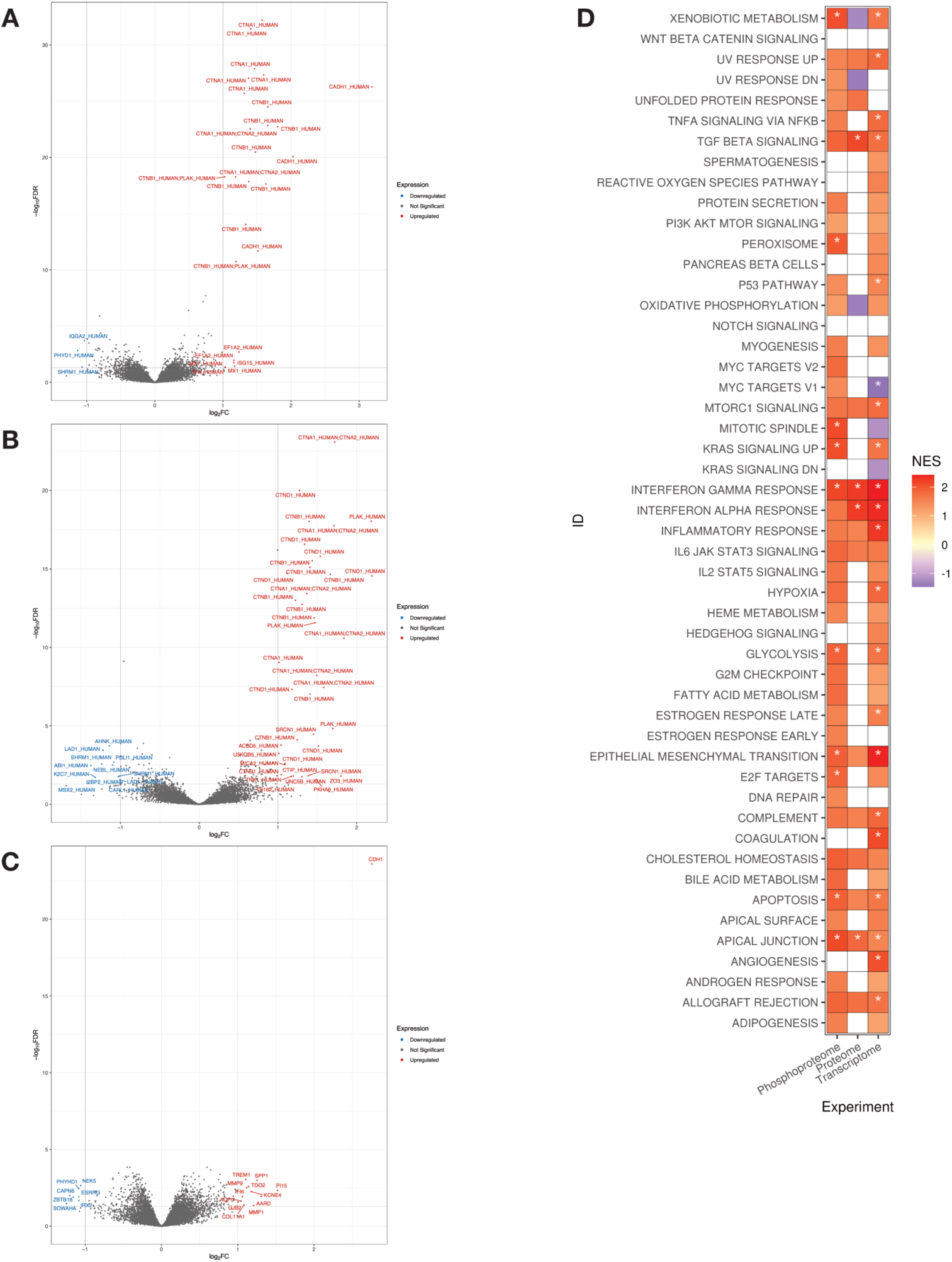
Differential features in NST versus ILC. A-C) Volcano plot of differentially abundant (A) proteins, (B) phosphopeptides, and (C) genes. D) Gene set enrichment analysis between NST and ILC across all three omics. FDR threshold was set at 0.25. Values with adjusted p-value < 0.01 are marked with an asterisk.

At the proteome level, comparing NST and ILC resulted in 29 differentially abundant protein groups (FDR<0.05, absolute log2 Fold Change > 1) (**Supplementary Table 1**). Of these, three were overexpressed in ILC, while the remaining 26 were overexpressed in NST. The most enriched protein in NST was Cadherin-1 (CADH1; adjusted p-value<5.5E-27, log2 Fold Change > 3). Moreover, several of the protein groups highlighted in this comparison have roles in cell adhesion, such as alpha and beta catenins (CTNA1, CTNA2, CTNB) as well as junction plakoglobins (PLAK).

At the phosphoproteome and transcriptome levels, a similar pattern is observed, where the most enriched features (FDR<0.05, absolute log2 Fold Change >1) in NST are related to cell adhesion (**Figure 2**).

Besides differential expression analysis, we conducted a Gene Set Enrichment Analysis (GSEA) of features found across all three omics layers (**Figure 2**). Cadherin-1 is listed as being part of three gene sets in the Hallmarks gene sets from MSigDB, namely *TGF_BETA_SIGNALLING*, *ESTROGEN_RESPONSE_LATE*, and *APICAL _JUNCTION*. Our results show that NST samples are significantly enriched (FDR<0.25) in all three pathways, with apical junction and TGF beta signaling pathways being enriched in all three omics layers, with at least two omics showing high significance (adjusted p-value < 0.01).

Our results thus corroborate that CADH1 and other features related to cell adhesion compose the main differences between NST and ILC, and the acquired multiomics data is in line with the biological characteristics of these two subtypes.

### Multiomics analysis reveals novel subtypes with distinct characteristics and survival

As our results captured known biological differences between the NST and ILC subtypes, we were interested in investigating whether the different omics would contribute to the identification of novel subtypes. To achieve that, we performed unsupervised consensus clustering at the proteome, phosphoproteome and transcriptome level.

As baseline, we first performed unsupervised consensus clustering at the proteome level for all available samples (177), resulting in 6 distinct clusters (**Figure 3A**). Nottingham histological grade (NHG), PAM50 subtype and the invasive histological subtype appear to show some correlation to the clusters. For instance, cluster 2 is dominated by grades 2 and 3 NSTs, whereas cluster 4 has, for the most part, grade 2 ILCs. Both clusters are dominated by the Luminal B subtype, whereas cluster 3 consists of mostly lower grade Luminal A cases.

**Figure 3.**
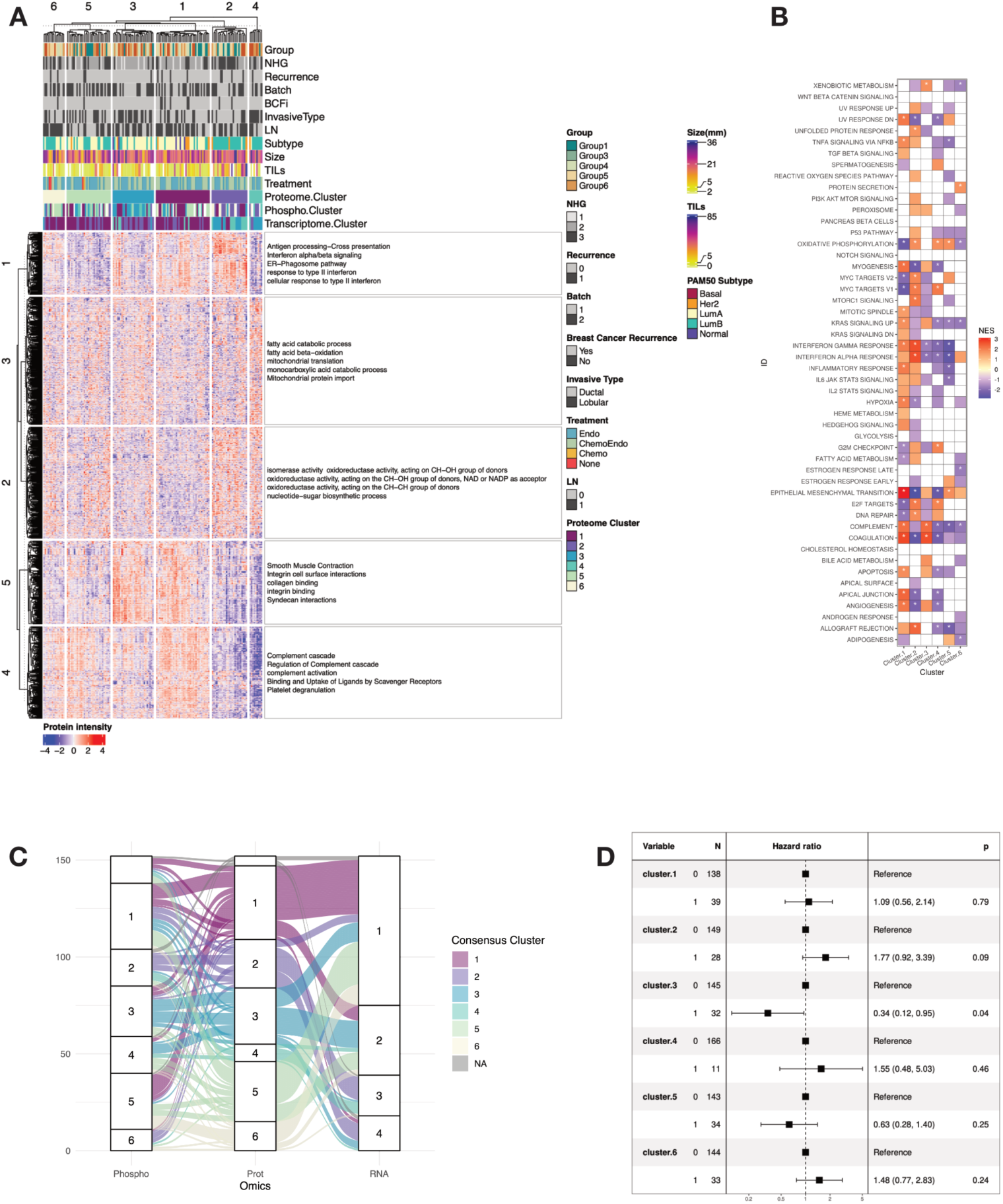
Multiomic identification of novel subtypes. A) Consensus clustering at the proteome level using the top 25% most variable proteins based on median absolute deviation. The proteins were further clustered in 5 distinct clusters based on K-means and the terms represent enrichment based on G:profiler. B) Gene set enrichment analysis in the proteome-defined clusters. FDR threshold was set at 0.25 and values with adjsuted p-value < 0.01 are marked with an asterisk. C) Alluvial plot demonstrating how samples in the proteome clusters get classified when consensus clustering is run at the phosphoproteome or transcriptome level. D) Forest plot of the clusters defined at the proteome level based on the univariable Cox regression model with recurrence free interval (RFI) as clinical outcome.

G:profiler ^24^ was used to add representative terms to each feature cluster. These results suggest that Cluster 2 is enriched in pathways related to interferon signaling and antigen processing and presentation, highlighting the potential role of an immune component in this cluster. This is further evidenced by the higher proportion of infiltrating lymphocytes as estimated by histological scoring in this cluster.

These annotations were further confirmed by performing differential expression analysis between the different clusters followed by GSEA at the proteome level (**Figure 3B**). Compared to the other proteome clusters, cluster 2 showed enrichment in, e.g., pathways related to interferon signaling, inflammatory response, MTORC1 and MYC signaling.

To further characterize the immune component of this cluster, we looked at the general immune cell marker PTPRC (CD45) in the proteome data, confirming significantly higher levels (FDR<0.05) in cluster 2 compared to the others, and further significantly lower levels in cluster 5. This protein also correlated (Spearman correlation coefficient 0.44) with histological scoring^18^ of lymphocyte cell infiltration percentages across the dataset. To gain insight about specific immune cell types, we performed transcriptome data deconvolution using CIBERSORTx, and significantly higher levels (FDR<0.05) of CD-4 and CD8 T-cells, as well as higher levels of monocytes and macrophages were found in cluster 2 compared to the others. The absolute CIBSERSORTx immune cell score was also higher in this cluster, in line with the PTPRC protein.

Performing consensus at the phosphoproteome level led to the identification of 5 clusters (**Supplementary Figure S1A**). A similar trend could be observed here compared to the proteome. Phosphoproteome clusters 2 and 5 were defined by being mostly composed of histological grade 3 tumors of Luminal B subtype, while the others were mixed.

In terms of GSEA, the pathways enriched in these two clusters show some overlap with those enriched in proteome cluster 2. The alluvial plot (**Figure 3C**) illustrates that, where samples belonging to proteome cluster 2 are mostly assigned to phosphoproteome clusters 2 and 5.

Finally, at the transcriptome level, 5 distinct clusters were defined (**Supplementary Figure S1B**). Associating the clusters with GSEA results also points to an enrichment of interferon signaling, inflammatory response and MTORC1 signaling in those clusters defined by higher histological grade, which can also be observed in the alluvial plot.

To further investigate these similarities, we applied a Cox proportional hazards model to compare the clusters in each omics layer and investigate if patterns in survival would follow the same trend (**Figure 3D**).

Looking at the forest plots for recurrence-free survival (RFS) for the proteome clusters, cluster 3 shows significant improved survival compared to the others (univariable p-value=0.04, HR = 0.34). Conversely, despite not being significant, cluster 2 shows overall worse survival (univariable p-value = 0.09, HR = 1.77). The GSEA results indicate opposite trends, where interferon and MTORC1 signaling appear to be downregulated in cluster 3 and upregulated in cluster 2. Interestingly, pathways associated with bile acid metabolism, coagulation and complement system are enriched in cluster 3.

### Identification of features associated with LN metastasis

As mentioned previously, the presence of tumor cells in lymph nodes is not only the earliest sign of dissemination, but it also constitutes an important prognostic marker in BC. For that reason, we were interested in finding features associated with LN metastasis

We first performed differential expression analysis between LN-positive and negative cases across both NST and ILC, excluding the subgroup of NST samples with a known distant recurrence event (Group 2). At an FDR cutoff of 0.1, two features (AKAP2 protein; PRC2A phosphopeptide, phosphorylation on Threonine 1347) passed. To detect systemic changes based on more subtle differences, we performed GSEA using the proteome, phosphoproteome and transcriptome data (**Figure 4A**). Several gene sets indicated similar trends across the omics layers, including interferon gamma and alpha response, complement, coagulation, and apical junction.

**Figure 4.**
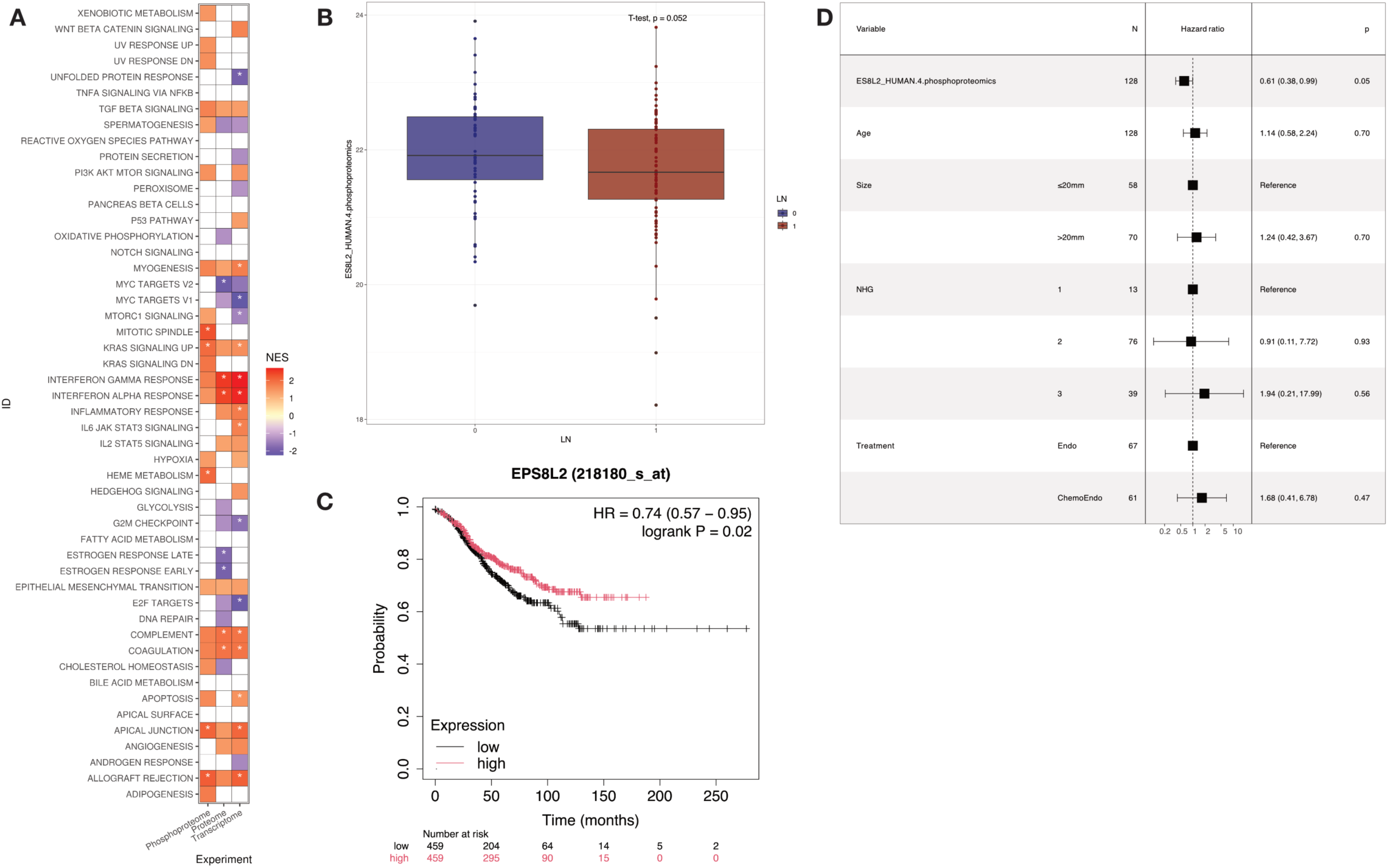
Features associated with lymph node involvement. A) Gene set enrichment analysis based on differentially expressed proteins,phosphopeptides and transcripts in LN-posive vs LN-negative (Group 2 excluded). B) Boxplot of normalized intensity values for ES8L2 S570p phosphopeptide. P-value based on Student’s t-test C) Kaplan-Meier plot for EPS8L2 from KM Plotter mRNA gene chip (see methods) based on ER-positive/HER2-negative LN-positive cases and using median RFS as survival outcome. D) Forest plot based multivariable Cox model using RFS for the ES8L2 S570p phosphopeptide.

As a way of finding more relevant features, throughout this study, feature selection was performed based on survival analysis. However, building a univariable Cox proportional hazards model with these two features did not result in significant association with RFS or overall survival (OS).

Considering that the results based on differential expression did not prove very insightful, we then investigated an alternative approach, based on integrating multiomics data. We employed Multi-Omics Factor Analysis (MOFA) ^25^ to the same samples used in the differential expression analysis. Akin to principal component analysis, MOFA allows for data decomposition in different (hidden) factors. The method allows for integration of multiple data modalities in an unsupervised fashion. Compared to other approaches, it allows for data reconstruction and association with clinical covariates, providing increased interpretability of the results ^25^.

Given the possibility to integrate different data modalities, besides the three omics datasets, we also included immune infiltration data based on two distinct deconvolution algorithms, CIBERSORTx ^26^ and EPIC ^27^. Both these algorithms are commonly used for deconvoluting transcriptome data, and they both belong to the reference-based category of deconvolution algorithms^28^. While initially developed for use with transcriptomics data, the algorithms are also capable of handling proteome data.

To select features associated with LN metastasis, we first selected latent factors from the MOFA model with a significant correlation to LN status and features correlated to these from each of the data layers, i.e., proteomics, phosphoproteomics, transcriptomics and deconvolution-based immune cell composition, resulting in a total of 289 unique features. These were individually run in a univariable Cox proportional hazards model to assess their association with RFS or OS. Those which did not violate the proportional hazards assumption were then further evaluated in a multivariable model (**Supplementary Table 2**). A total of 14 features passed univariable test, and 3 passed the multivariable test, but only one (ES8L2 phosphopeptide, phosphorylation on Serine 570, multivariable p-value = 0.05, hazard ratio 0.61) proved to be significant in both (**Figure 4D**). Of note, **Figure 4B** depicts a boxplot of the normalized phosphopeptide intensities between the two LN groups. The two means were compared via Student’s t-test, showcasing marginally significant (p-value = 0.052) results supporting a decrease in abundance for this peptide in connection to LN involvement.

Repeating the same process with OS as the survival endpoint resulted in 58 features passing the univariable test. In the multivariable setting, 4 features were considered significant, but none were significant on both tests (**Supplementary Table 2**).

To validate these findings, we further evaluated ES8L2 in the Human Protein Atlas (HPA)^29,30^, Kaplan-Meier Plotter^31,32^ and with RNAseq data available from other samples belonging to the SCAN-B cohort ^18,20,29,32^, as no additional phosphoproteome data were available for this purpose.

According to UniProt, the ES8L2 protein corresponds to the epidermal growth factor receptor kinase substrate 8-like protein 2, and it acts in the stimulation of Ras-bound guanine exchange activity of SOS1 (son of sevenless homolog 1) and has a possible role in remodeling of the actin cytoskeleton.

In the HPA BC pathology, ES8L2 is not listed as prognostic in BC, as only features with a p-value lower than 0.001 receive such denomination. The Log-Rank p-value based on the best transcript expression cutoff, i.e., the FPKM value which yields maximal difference with regards to survival between the two groups was 0.014, with the 5-year survival being 84% for the high group and 80% for the low group, i.e., it appears to be associated with improved survival.

In the Kaplan-Meier plotter, at the mRNA level, selecting for ER-positive/HER2-negative samples and using RFS as the survival endpoint with the cutoff at the median resulted in a p-value of 0.39 and hazard ratio of 0.93. However, when stratifying cases further based on LN status, a significant trend was detected in LN-positive cases (p-value 0.02, HR = 0.74) (**Figure 4C**).

Finally, when using the transcriptome data available from other SCAN-B samples, a similar trend was observed in RFS. It was considered significant in both univariable and multivariable tests for mixed cases, i.e., both LN statuses in conjunction (number of cases = 3355, univariable p-value = 0.02, HR = 0.86, multivariable p-value = 0.003, HR = 0.082), and when LN-negative cases were analyzed separately (number of cases = 2097, univariable p-value = 0.002, HR = 0.81, multivariable p-value = 0.014, HR = 0.79). It was not deemed significant when LN-positive cases were analyzed separately. When using OS as the survival endpoint, the ES8L2 transcript was significant in univariable Cox models for all samples and LN-negative cases separately, but it did not pass the multivariable tests. Of note, despite significance, the calculated hazard ratios had opposite trends when comparing RFS and OS, i.e., the transcript was associated with improved RFS and worse OS.

### Identification of features associated with distant metastasis

Metastatic disease is the main cause of cancer-related deaths^11^. Nevertheless, the location of metastasis also holds prognostic and diagnostic value, with loco-regional metastases being considered curable stage III disease, and distant metastases constituting stage IV disease, at which stage palliative care is generally used^33^.

Cancer cells must travel different distances in order to reach local lymph nodes and distant sites, which ultimately exerts different selection pressures on them, culminating in distinct molecular characteristics^33^.

With this aspect in mind, our focus was on identifying features linked to the presence of distant metastasis. To separate the two metastatic events, we focused on a subset of 61 LN-negative NST samples, Group 1 and Group 2, of which the later was selected for having a distant recurrence event.

Initial differential expression analysis identified a total of five distinct features at FDR <10%, a protein group composed of heat shock protein 90 alpha and beta (H90B3;HS902;HS90A;HS90B, adj. p-value=0.054), and four phosphopeptides, namely serine/threonine-protein kinase SIK2 (SIK2, phosphorylation residue serine 534, adjusted p-value = 0.08), RelA-associated inhibitor (IASPP, phosphorylation residue threonine 173, adjusted p-value = 0.08), Neuron navigator 1 (NAV1, phosphorylation residue serine 391, adjusted p-value = 0.08), and Lamin-B2 (LMNB2, phosphorylation residue threonine 34, adjusted p-value = 0.09).

The results from this step were also used in GSEA to assess potential pathways involved in the two groups, despite no further individual proteins being significant after p-value adjustment for multiple testing (**Figure 5A**). Pathways associated with estrogen response – both early and late – and epithelial mesenchymal transition appear enriched in the group with no distant recurrence event. On the other hand, pathways involved in G2M checkpoint, E2F targets, MYC targets, MTORC1 signaling and interferon alpha and gamma response, show some enrichment in the group with distant recurrence. To further characterize differences and similarities between lymph node and distant metastases, we utilized the phosphoproteome data to search for common and distinct signatures using PTM signature enrichment analysis^34^ (**Figure 5B**). Interestingly, the general pattern of enrichment score directions were similar between the two groups when compared to the lymph node negative samples for signatures with significant enrichment in any of the comparisons (FDR<0.01), although with different significance levels. However, only the perturbation signature PSP_PHORBOL_ESTER was significantly enriched in the control group vs the two metastasis groups. The kinase signatures iKiP_STK4 and iKiP_NEK6 were significantly enriched in the lymph node metastasis group, while the kinase signature iKIP_CDK2_CCNA2 was significantly enriched in the distant metastasis group.

**Figure 5.**
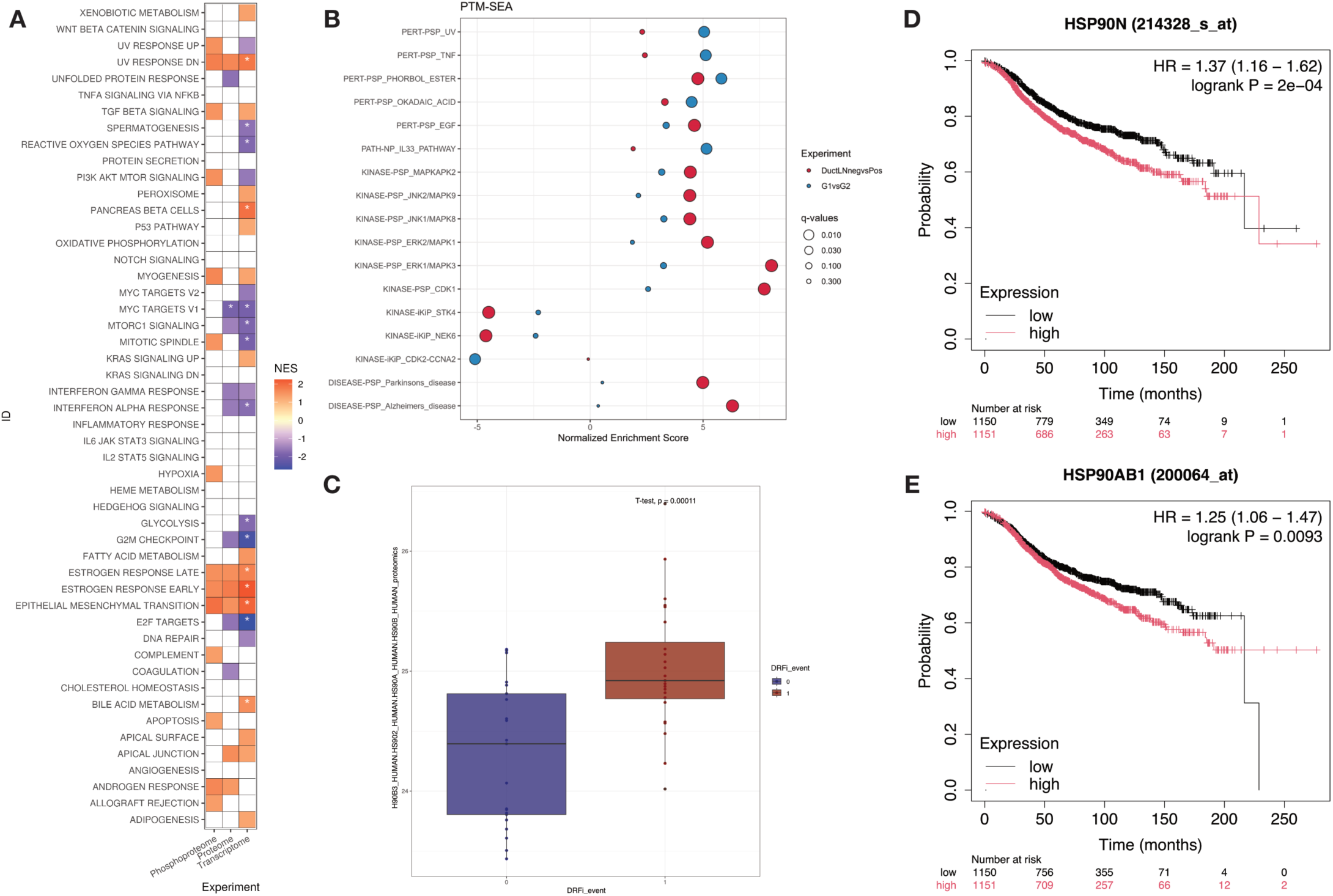
Features associated with distant metastasis. A) Gene set enrichment analysis based on differentially expressed proteins,phosphopeptides and transcripts in Group 1 vs Group 2. FDR threshold was set at 0.25 and features with adjusted p-value < 0.01 are marked with an asterisk. B) Plot based on PTM-SEA analysis of phosphopeptides in either LN-negative vs LN-positive (red) or Group1 vs Group 2 (blue). C) Boxplot of normalized intensity values for the Heat Shock Protein 90 protein group. P-value based on Student’s t-test D-E) Kaplan-Meier plot for HSP90AA1 (D) or HSP90AB1 (E) from KM Plotter mRNA gene chip (see methods) based on ER-positive/HER2-negative cases and using median RFS as survival outcome.

Both univariable and multivariable cox proportional hazards models were used to evaluate the association of these differentially abundant features with survival. Since the event separating the two groups is distant recurrence, Distant Recurrence Free Survival (DRFS) was used as the survival outcome instead of RFS, given that it is more specific.

With DRFS as the survival outcome, all five features were deemed significant in both univariable and multivariable models. In general, the protein group H90B3;HS902;HS90A;HS90B (univariable p-value < 0.001, HR = 2.25, multivariable p-value = 0.006, HR = 2.10), the SIK2 S534p (univariable p-value < 0.001, HR = 2.44, multivariable p-value = 0.003, HR = 2.25), and NAV1 S391p (univariable p-value < 0.001, HR = 2.04, multivariable p-value = 0.001, HR = 2.17) were associated with worse DRFS survival, while LMNB2 T34p (univariable p-value < 0.001, HR = 0.55, multivariable p-value = 0.02, HR = 0.60) and IASPP T173p (univariable p-value = 0.002, HR = 0.50, multivariable p-value = 0.02, HR = 0.55) were associated with improved DRFS survival (**Supplementary Table 3**).

In terms of OS, the Heat Shock Protein 90 protein group, and phosphopeptides from LMNB2 and SIK2 were significant in a univariable model following the same trends observed with DRFS. From these three, H90B3;HS902;HS90A;HS90B and SIK2 were also significant in a multivariable model for OS (**Supplementary Table 3**).

In addition to the differential expression analysis, we built a MOFA model including immune deconvolution data from CIBERSORTx and EPIC in addition to the three omics layers described across the subset of 61 samples mentioned previously in this section. The same strategy as for assessing feature LN involvement was used, with feature selection based on association with latent variables correlating with distant metastasis, followed by analysis in univariable and multivariable Cox proportional hazards models.

Across the different omics and cell types derived from the deconvolution algorithms, a total of 53 features were selected. Of these, 50 passed the proportional hazards assumption test. **Supplementary Table 4** contains these selected features, and the statistics derived from univariable and multivariable Cox models. A total of 5 features had p-value lower than 0.05 in both models with DRFS as the survival endpoint.

Analyzing the same 53 features with OS as the survival endpoint resulted in 49 features passing the proportional hazards assumption (**Supplementary Table 4**). Of these, only one was considered significant on both univariable and multivariable analysis, namely RBM39 (univariable p-value = 0.005, HR = 2.30, multivariable p-value = 0.04, HR = 2.28).

External validation of these findings – performed in the HPA, Kaplan-Meier Plotter and with the transcriptome data from other SCAN-B samples^20^ – was in line with what was conducted for the feature associated with lymph node metastasis.

Starting with the H90B3;HS902;HS90A;HS90B protein group, the genes HSP90AA1 and HSP90AB1 were searched in the HPA. Both are considered significant with higher expression being linked to worse survival. Moreover, HSP90AA1 is also listed as prognostic in BC (p-value = 0.00056). Searching for the same two genes in KM Plotter within ER-positive/HER-negative cases and using DRFS and OS as the survival outcomes resulted in only HSP90AB1 being significant in LN-negative cases for DRFS (p-value = 0.0058, HR = 1.81). Of note, if using RFS as the survival outcome, both genes are significant (**Figure 5D, E**) in both mixed LN status and LN-negative cases.

The analysis of these two genes using transcriptome data from other SCAN-B samples did not result in any significant observation, although the normalized intensity of the corresponding protein group in our analysis proved highly significant (**Figure 5C**).

Among corresponding genes from the four phosphopeptides identified via differential expression analysis, only LMNB2 was considered significant in both the HPA (p-value = 0.039) and KM Plotter (DRFS p-value = 0.0088, HR = 1.49), in both cases with higher expression being linked to worse outcome. However, employing the complementary SCAN-B validation, LMNB2 was the only gene among these which showed some degree of significance, albeit only in LN-negative cases and univariable analysis (number of cases = 2098, p-value = 0.003, HR = 1.41).

Moving on to the features identified via MOFA, in the HPA, all but MFGM were considered significant. The corresponding genes from the protein groups TSP1;TSP2 (TSP1 p-value = 0.0017, TSP2 p-value = 0.029) and RBM39 (p-value = 0.014) were associated with worse survival, while those from MYH10.MYH11.MYH9 (only MYH10 was significant, p-value = 0.047) and KCND3 (p-value = 0.011) were linked with improved survival. Subsequent validation in the KM Plotter of the same genes resulted in two significant hits with DRFS as the outcome, namely MYH10.MYH11.MYH9 (only MYH11 was significant, p-value = 0.00058, HR = 0.59) and KCND3 (p-value = 0.0035, HR = 1.55). Finally, in the SCAN-B validation, the results were mixed, with some features only being significant in univariable or multivariable models. Within LN-negative cases, however, the corresponding genes for RBM39 (univariable p-value = 0.012, HR = 0.71, multivariable p-value = 0.005, HR = 0.69) and MFGM (univariable p-value = 0.037, HR = 1.36, multivariable p-value = 0.01, HR = 1.42) were evaluated as significant in both univariable and multivariable analysis.

## Discussion

In the present study, we generated a multiomic profile of metastatic processes in ER-positive/HER2-negative BC, combining transcriptomics, proteomics, phosphoproteomics and immune infiltration estimates from the same original sample material. This was achieved by utilizing flowthroughs collected via standard procedures as integral part of the SCAN-B cohort sample processing pipeline. Building on a previously developed semi-automated protocol ^19^, we acquired phosphoproteome DIA data in addition to the already standard proteomic DIA data. The proteome and phosphoproteome data were then integrated with already published transcriptome data, generating the most comprehensive dataset of its kind in ER-positive/HER2-negative BC. Moreover, new to this study was the integration of such data with immune infiltration estimates generated by two deconvolution algorithms, EPIC and CibersortX.

Our multiomic analysis enabled the identification of 6 distinct clusters based by consensus clustering at the proteome level. While all clusters possess distinct characteristics in terms of enriched hallmark pathways and survival, two were of particular interest, namely clusters 2 and 3. In our univariable survival analysis, cluster 3 was significantly associated with improved RFS, while cluster 2 had the opposite trend, although not significant (**Figure 3D**).

Together with the heatmap shown in **Figure 3A**, which showed higher grade luminal B tumors with higher immune infiltration for Cluster 2, the GSEA results highlighted pathways associated with e.g., interferon (IFN) signaling (i.e., interferon alpha response and interferon gamma response) as well as MYC targets, IL6 JAK STAT3 signaling, DNA repair and allograft rejection. Of note, the identification of an immune hot cluster is in line with the results reported in Asleh, K. et. al. (2022)^35^, although our findings concern ER-positive/HER2-negative samples exclusively.

In broad terms, IFNs are cytokines produced primarily in antiviral response ^36^, although their functions related to immunity and cancer have also been documented ^36–38^. IFNs are broadly classified in three types, type I, consisted of IFN-alpha and IFN-beta, type II (IFN-gamma) and type III (IFN-lambda). While types I and III are produced by different cells, type II is mainly produced by T cells and NK cells, underscoring its role in immune responses ^36^.

The use of IFNs as therapeutic agents was also suggested in the past ^36^. For instance, the use of IFN-beta in combination with tamoxifen was an early strategy to circumvent hormonal therapy resistance in ER-positive BC ^36,39^. However, the complexity of IFN signaling in conjunction with substantial toxicity ultimately limited their use ^36^.

Since then, it has been debated that IFN has a dual role in the tumor microenvironment (TME), being able to exert both anti-tumor and pro-tumor functions ^36,40^. For example, it has been reported that chronic exposure to IFN-gamma would induce the expression of inhibitory factors, ultimately leading to immune exhaustion and evasion due to the immunosuppressive TME ^36,41,42^.

When it comes to the role of the immune system in BC, distinct immune biology can be observed in different BC subtypes, with ER-negative tumors, i.e., HER2-positive and triple negative tumors (TNBC) typically having increased immune infiltration – defined by tumor infiltrating lymphocytes (TILs) – in comparison to ER-positive tumors, making those more immunogenic and, by consequence, targetable by immunotherapy ^43,44^. Still, our results suggest that there could be subsets of ER-positive/HER2-negative tumors with potentially actionable immune profiles.

Denkert and colleagues (2018)^45^ analyzed data from 3771 BC patients treated with neoadjuvant chemotherapy originated from six randomized, multicenter clinical trials with the goal of assessing the link between the presence of TILs and response to treatment and survival, as well as the differences between luminal/HER2-negative tumors and TNBC. As expected, the percentage of cases with high TILs in TNBC and HER2-positive BC was higher than in luminal/HER2-negative cases^45^. When assessing the prognostic value of TILs in overall survival and disease-free survival by means of logistic regression, the authors observed that, unsurprisingly, higher TILs contributed to longer disease-free survival and overall survival in TNBC. In luminal/HER2-negative cases, a negative association was seen in overall survival, i.e., low TILs were significantly associated with longer overall survival^45^. Similar trends were observed when TILs were stratified into low, intermediate and high groups, where higher TILs were associated with both decreased overall survival and disease-free survival in luminal/HER2-negative cases^45^.

Of note, contrary to these findings, a study conducted by Lundgren and colleagues (2020) showed that improved prognosis, i.e., Breast Cancer-Free interval (BCFi), was seen in cases with high TILs infiltration, irrespective of subtype, seen in both univariable and multivariable analyses. For ER-positive/HER2-negative cases alone, multivariable analysis was significant for both BCFi and OS, despite non-significant univariable results. The study included a randomized trial of premenopausal patients of different BC subtypes with over 30 years of follow-up data^44^.

The predictive value of TILs infiltration in ER-positive cases with 2 years of adjuvant tamoxifen therapy (TAM) was also assessed. Here, it was shown that TAM improved BC survival (BCFi) for the groups with low and intermediate TILs, although the predictive value of TILs could not be confirmed^44^.

Since the immune component refers to different cell types, Denkert and colleagues also investigated samples from the Metabric database ^45,46^ as well as TNBC and luminal/HER2-negative cases. Based on available mRNA expression data, a microenvironment-cell-populations (MCP) counter method was applied to estimate the abundance of different immune cell populations ^45^. They observed that while most immune cells in TNBC were associated with better prognosis, in luminal/HER2-negative cases, only B cells and myeloid-derived dendritic cells (DCs) were associated with improved prognosis. In contrast, monocytes or macrophages were linked to poor prognosis ^45^.

These findings align with our own results, since Cluster 2 has the highest immune infiltration, consisted mainly of increased T cells – CD8+, CD4+ and regulatory T cells (Tregs) – as well as macrophages (**Supplementary Figure S2**). Interestingly, although M0-like and M2-like macrophages are comparable across the different clusters, Cluster 2 also shows increased presence of M1-like macrophages (**Supplementary Figure S2**).

Cytotoxic T cells are the main producers of IFN-gamma, which is also responsible for the upregulation in the expression of inhibitory factors such as programmed cell death protein ligand 1 (PD-L1) in macrophages and tumor cells with chronic exposure, resulting in a tumor suppressive environment ^36^. This profile with increased IFN signalling and possible T cell exhaustion could potentially be the target or immune checkpoint blockade (ICB) therapy ^36,47^.

The other cluster that stood out in our analysis was Cluster 3, this time for a significant association with improved RFS. In terms of the variables included in the heatmap presented in **Figure 3A** and the immune infiltration results, this cluster is primarily characterized by lower grade tumors of the luminal A subtype.

Turning to the list of differentially expressed features and the GSEA analysis, this cluster is associated with significant positive enrichment in pathways related to complement, coagulation and xenobiotic metabolism, and significant negative enrichment in pathways linked to IFN signaling. The list of differentially abundant proteins is dominated by apopoliproteins (APOs), with apopoliprotein A1 being the most significant differentially abundant protein (APOA1, adj. p-value = 5.12E-10, log2 FC = 1.07).

In general, APOs are proteins that bind to lipids and can act as ligands for cell surface receptors, enzymatic cofactors and lipid carriers, where they are typically divided based on density into very-low-density lipoprotein (VLDL), low-density lipoprotein (LDL), intermediate-density lipoprotein (IDL) and high-density lipoprotein (HDL)^48^.

The most abundant lipoprotein is HDL, and it has been associated with the regulation of different immune functions, as well as having anti-inflammatory, anti-oxidative and anti-apoptotic properties ^49^. Although it has also been associated with the risk of different cancers, it is unclear whether HDL itself or its main constituent APOA1 is the main responsible for exerting these roles ^49^.

Pedersen et. al. (2020) ^49^ tested the hypothesis that low HDL is associated with increased risk of cancer. For this purpose, individuals from two independent Danish population-based cohorts (16,728 individuals in total) were followed for up to 25 years and the risk of different cancer types was investigated ^49^. The authors observed an increased risk of BC with low levels of HDL/APOA1, although this was not considered significant upon correction for multiple hypotheses ^49^. They further discussed possible mechanisms mediating this protective effect and highlighted the ability of HDL to regulate immune response, inflammation and apoptosis ^48,49^, as well as a possible inhibition in the proliferation of hematopoietic stem cells ^49,50^, which would fall in line with the previously discussed associations between worse prognosis in highly infiltrated luminal/HER2-negative tumors.

Our analysis of multiomic features associated with lymph node metastasis led to the identification of a phosphopeptide belonging to the EPS8L2 protein. EPS8L proteins constitute a family of EPS8-related proteins which is composed of three members, EPS8L1, L2 and L3 ^51,52^. The EPS8 protein is essential for activation of Rac via the formation of a tricomplex (EPS8-SOS1-ABI1) ^51,52^.

In the work of Offenhäuser et al (2004) ^51^, the authors discuss that despite the essential role of EPS8, EPS8 null mice lack an associated phenotype, with the most plausible explanation being the existence of redundant functions ^51^. The family of EPS8L proteins was further analyzed in their capacity to form the tricomplex, interact with actin and recover EPS8 activity in EPS8 −/− fibroblasts derived from EPS8 knockout mice ^51^. The authors concluded that the expression pattern of EPS8 and EPS8L2 were overlapping, leading to functional redundancy at the protein level ^51,52^. Moreover, in terms of Rac activation via the tricomplex, EPS8L2 was equally efficient as EPS8 itself^51^.

These findings and functional redundancy led to the investigation of potential roles of EPS8L2 and EPS8. In their review on the effects and mechanisms of EPS8 in malignant tumors, Luo et. al. ^52^ discuss the expression of EPS8 in different cancers (both solid tumors and hematological malignancies) and highlight the potential use of EPS8 as a biomarker or target ^52^.

EPS8 is part of a variety of signaling processes, including epidermal growth factor receptor (EGFR) transduction, actin binding and regulation of cell cycle and cell proliferation ^52^, where it is typically seen as a marker of poor prognosis and generally associated with tumorigenesis, proliferation, migration, metastasis, chemoresistance ^52,53^.

Contrary to these findings, He et. al. ^53^ demonstrated that EPS8 can induce maturation of dendritic cells, T-cell proliferation and cytotoxicity in vitro, with significant increases in the expression of major histocompability complex class II (MHC-II), CD80, CD86, interleukin 12 (IL-12) and increased IFN-gamma secretion^53^.

A murine breast cancer model, 4T1, was used to investigate the effect of an EPS8 vaccine^53^. Even though no mice remained tumor free, immunization with EPS8 vaccine inhibited tumor growth considerably and led to longer survival with subsequent injection of subcutaneous 4T1 cells ^53^. A reduction in the percentage of Tregs was also observed ^53^.

Considering our own findings, where EPS8L2 S570p is linked to improved RFS and appears to have lower expression in LN-positive cases, a possible mechanism involving a protective role of EPS8L2 could be in reducing the population of Tregs and mitigating the potential T-cell exhaustion previously mentioned. Similarly, given the increased IFN-gamma expression observed by He et. al. ^53^, it is possible that ICB therapy could be applied in some cases. However, these mechanisms and uses need further investigation. Finally, in our validation results, EPS8L2 showed different associations with RFS and OS, which could, at least in part, be explained by functional redundancy with EPS8 and the intricate signaling pathways associated with it, besides possible differences resulted from the validation being done at the transcriptome level.

Continuing with the findings from our multiomic analysis of features associated with distant metastasis, one of the features highlighted were a protein group composed of different isoforms of the Heat Shock Protein 90 (HSP90).

In general, Heat Shock Proteins (Hsps) are highly conserved, and their expression is upregulated in response to stress, be it physical, chemical or in pathological processes, including carcinogenesis ^54,55^. In mammals, Hsps are divided into six different families based on their molecular size, with the HSP90 family being encoded by *HSPA* genes ^54^. In relation to their role in cancer, the expression of different Hsps has been shown to be increased in several different tumors, including BC ^54^.

The HSP90 proteins are constituted by three domains, an N-terminal domain, a C-terminal domain and a middle domain ^54,55^. The HSP90 family is crucial for their chaperone activity, assisting in the correct folding of its client proteins and inducing protein degradation where appropriate ^55^. To date, over 400 different client proteins have been identified, several of which are essential for cancer cell proliferation ^54,55^. In BC, expression of HSP90 is typically associated with a worse prognosis ^54,56,57^.

Considering that HSP90 is typically overexpressed in BC and that it is associated with worse prognosis, its inhibition could prove beneficial. In fact, different HSP90 inhibitors have been proposed and evaluated in different clinical trials ^54,55^. Li et. al. ^55^ reported a total of 18 HSP90 inhibitors divided in five different categories associated with their chemical structures.

Historically, most HSP90 inhibitors targeted the N-terminus domain. However, these typically lead to the development of toxicity and resistance by means of upregulation of other Hsps, resulting in a process known as the heat shock response ^54,55^. Targeting the C-terminal, on the other hand, appears to partially mitigate the issue ^55^.

On the other hand, it has been shown that the HSP90 family is composed of four different isoforms, namely HSP90-alpha, HSP90-beta, GRP94 and TRAP-1 ^55^. Of these four, the alpha and beta isoforms are the most abundant ^55^. For that reason, the use of isoform specific inhibitors that target these more abundant isoforms, such as SNX-0723 and TAS-116, would be the optimal choices due to increased selectivity and reduced toxicity, in monotherapy or combination therapy scenarios ^55,58,59^.

In addition to the use of inhibitors, Zhu et. al. ^60^ developed a risk signature for BC based on markers involved in immunogenic cell death (ICD), a process which involves the transformation of tumor cells from non-immunogenic to an immunogenic phenotype given a death stimulus ^60^. Among the different markers identified, the authors highlight the *HSP90AA1* gene as that with the highest risk factor and further demonstrate that knockdown of *HSP90AA1* with siRNA resulted in a significant reduction in BC cell migration and invasion ^60^, underscoring the potential value as a biomarker of metastasis. Our results both from the multiomic discovery cohort and in the external validations agrees with the potential usage of this protein or transcript as a predictive biomarker, especially in LN-negative cases.

## Conclusions

In the present study, we present the most comprehensive multiomic dataset of metastatic processes in ER-positive/HER2-negative BC clinical samples. This was achieved by further developing a workflow and combining LC-MS/MS-based proteome and phosphoproteome data, and immune infiltration estimates with transcriptome data from tumor biopsies, followed by integrative omics analysis methods such as consensus clustering and MOFA. We identified possible subtypes with differential survival, highlighting a possible immune exhausted phenotype. Finally, we identified potential markers of LN involvement and distant metastasis which are part of several important signaling pathways that are targets of active clinical developments. Further investigation of their potential predictive and prognostic roles of these markers could be crucial in the context of personalized medicine in ER-positive/HER2-negative BC.

## Methods

### Ethics approval and consent to participate

Included patients were enrolled in the Sweden Cancerome Analysis Network – Breast (SCAN-B) study (ClinicalTrials.gov ID NCT02306096) (PMID:25722745,29341157) approved by the Regional Ethical Review Board in Lund, Sweden (registration numbers 2009/658, 2010/383, 2012/58, 2013/459, 2014/521, 2015/277, 2016/541, 2016/742, 2016/944, 2018/267 and the Swedish Ethical Review Authority (registration numbers 2019-01252, 2024-02040-02). All patients provided written informed consent prior to enrolment and all analyses were performed in accordance with patient consent and ethical regulations and decisions.

### Sample Selection and Study Design

All samples used in the present study are from the Sweden Cancerome Analysis Network – Breast (SCAN-B) cohort^18,20^. A total of 182 samples were obtained in collaboration with the SCAN-B Steering Committee and the Division of Oncology, Department of Clinical Sciences Lund, Lund University.

The samples correspond to the flowthrough resulted from RNA and DNA extraction performed in tumor biopsies using the Qiagen All Prep kit^18^. This material contains the protein fraction, which was used in the present study for proteome and phosphoproteome data acquisition.

The clinical specimens were selected from the early-stage IBC cohort defined in ^20^. In summary, this cohort constitutes of non-redundant IBC cases representative of the background IBC population in the SCAN-B catchment area ^20^. From this cohort, samples were selected to only comprise ER-positive, HER2-negative tumors collected between 2010 and 2016 to allow for at least 5-year follow-up data to be available and standardize treatment. The samples were then further filtered based on the interquartile range of size with the objective of standardizing size across samples, given it is highly correlated with aggressiveness and metastatic spread.

The aim of the study was to profile the lymph node involvement and distant metastasis in BC. With that in mind, the samples were then split into 6 experimental groups, which can be observed in **Figure 1**. First, samples were divided according to the two histological subtypes, i.e., 120 NSTs and 62 ILCs. These groups were further stratified in LN-positive and LN-negative, resulting in an average of 60 samples per group for NST and 31 per group within ILC.

Given that NST is much more prevalent than ILC, the LN-negative samples were further stratified according to the absence or presence of distant recurrences, Group 1 and Group 2, respectively. The LN-positive samples were divided according to nodal burden, with samples classified as 1to3 being placed in Group 3, and 4toX samples composing Group 4. For ILC samples, Group 5 corresponds to LN-negative and Group 6 to LN-positive.

In summary, an average of 30 samples per group was selected, preferring samples belonging to the Follow-up Cohort^20^, collected between 2010 and 2012 and completing the group with samples up until 2016. A Shapiro-Wilk test of normality was performed for the size of all defined groups. Apart from Group 6, all other groups had normal size distributions. No further corrections were made for Group 6, given the very limited number of samples available to satisfy the selection criteria.

### Estimation of Protein Concentration

The protein concentration in the samples was estimated based on regression analysis described in^19^. Given the poor correlation between protein and RNA concentration in the lower range, a cutoff of 200 ng/µL was stipulated and applied as a selection criterium in the present study.

### Sample Preparation

A total of 100 µg of each sample was transferred to KingFisher 96 deep-well microtiter plates (Thermo Fisher Scientific, Waltham, MA, USA). The volumes were normalized to 200 µL using MS-grade water.

#### Reduction and Alkylation

The proteins were then reduced and alkylated for 1 hour at room temperature to a final concentration of 5 mM Tris(2-carboxyethyl)phosphine hydrochloride (TCEP-HCl) and 10 mM 2-Chloroacetamide (2-CAA), using 100 mM stocks from each.

#### Automated Digestion via Protein Aggregation Capture

Protein Aggregation Capture (PAC) was performed for protein digestion. The protocol was automated using a KingFisher Flex robot (Thermo Fisher Scientific). Briefly, the tip comb was stored in position #1. In position #2, the plate containing the reduced and alkylated samples was added. To it, 10 µL MagReSyn® Hydroxyl beads (Resyn Biosciences, Edenvale, Gauteng, South Africa), corresponding to a bead-to-protein ratio of 2:1 (w/w), was added, followed by addition of Acetonitrile (ACN) to a final concentration of 70%. The beads were washed three times in 95% ACN in plates #3, #4 and #5, followed by two washes with 70% ethanol (plates #6 and #7). Finally, the proteins were digested overnight in 50 mM Ammonium Bicarbonate in the presence of Lys-C (1:500 enzyme/protein, w/w) and Trypsin (1:250 enzyme/protein, w/w). Digestion took place at 37°C with intermittent agitation, i.e., intervals of 15 seconds of agitation followed by 135 seconds without agitation. Protease activity was quenched by adding trifluoroacetic acid (TFA) to a final concentration of 1%. The resulting peptide solutions were stored at −80°C until desalting.

The resulting peptide solutions were dried on a SpeedVac Vacuum Concentrator (ThermoFisher Scientific) and stored at −80°C until desalting.

#### Desalting

The peptide mixtures were desalted on Sep-Pak 40 mg tC18 96-well plates (Waters Corporation, Milford, MA, USA). The columns were first conditioned with 100% ACN using a centrifuge at 100G for 1 minute, and the flowthrough was discarded. Then, equilibration was performed three times using 0.1% TFA in water. After each step, the flowthrough was discarded. The acidified peptide samples were loaded and passed through the columns at 80G on the centrifuge. At this stage, the flowthrough was collected, and the loading step was repeated a total of three times. The columns were washed three times with 0.1% TFA and the cleaned peptides were eluted in two steps, with 40% ACN and 70% ACN, respectively. A small aliquot of the cleaned peptide mixtures was taken for full proteome analysis. This aliquot was dried on a SpeedVac Vacuum Concentrator (ThermoFisher Scientific) and kept at −80°C until LC-MS/MS analysis. The remaining eluate was stored at −80°C until phosphopeptide enrichment was conducted.

As a result of a technical error during the drying step after phosphopeptide enrichment, the samples were lost. The protocol was run for a second time with minor differences from the steps mentioned above. Specifically, the desalting of peptides was performed using Oasis Prime HLB 10 mg 96-well plates (Waters Corporation). For this round of desalting, columns were conditioned with ACN in a centrifuge at 100G for 1 minute. Unless otherwise specified, the flowthroughs after each centrifugation step were discarded. Equilibration was performed using 0.1% TFA at the same speed setting. The acidified samples were loaded, and at this stage the flowthroughs were collected, and this step was repeated two additional times. Washing was then performed using 0.1% TFA, and the peptides were eluted in two steps using 60% ACN, 5% TFA and 0.1M GA. The samples were stored at −80°C until ready for phosphopeptide enrichment.

#### Automated Phosphopeptide Enrichment via Immobilized Metal Ion Affinity Chromatography (IMAC) Beads

Automated Phosphopeptide enrichment was automated using the KingFisher Flex robot (ThermoFisher Scientific) and based on^61^. The tip comb was stored in position #1. In position #2, 10 µL MagResyn® Zr-IMAC HP beads (Resyn Biosciences) were added in addition to 200 µL binding solution, which consisted of 80% ACN, 5% TFA, 0.1 M glycolic acid (GA). The beads were then washed once in binding solvent in position #3. In position #4,200 µL 5% TFA, 0.1M GA in ACN was added and mixed with the desalted peptides. In position #5, the beads were washed in binding solution, followed by two consecutive washes in positions #6 and #7 in 80% ACN, 1% TFA, and 10% ACN, 0.2% TFA, respectively. Elution was performed in 1% ammonia solution. A second round of enrichment was performed by looping over the protocol once without adding new beads. Enriched peptides were then acidified with 10% TFA, transferred to PCR plates and dried on a SpeecVac.

The current protocol was devised to allow for parallel processing of samples for acquisition of full proteome and phosphoproteome data. Unfortunately, due to a technical error during the drying step after phosphopeptide enrichment, the samples were lost at this stage. The protocol was rerun with minor changes. Specifically, the desalting step for the second run was performed in Oasis Prime HLB 10 mg 96-well plates (Waters Corporation).

#### Evotip Loading

For full proteome analysis, sample concentration was determined using a Nanodrop ND-1000 Spectrophotometer (ThermoFisher Scientific). Based on the concentration results, approximately 600 ng of peptides were loaded onto Evotip Pure tips (EvoSep Biosystems, Odense, Denmark) with minor modifications from the manufacturer’s instructions. Briefly, the equilibration step and the washing step were repeated an additional time, and 300 µL of solvent A (0.1% formic acid in water) was added to the tips in the last step, instead of the recommended 100 µL.

For loading of the enriched phosphopeptides, the entire eluate resulting from the enrichment step was loaded onto the Evotip Pure tips (EvoSep Biosystems). The modifications from the manufacturer’s protocol were also kept.

### LC-MS/MS

LC-MS/MS was performed on an Evosep One LC system (EvoSep Biosystems) coupled to an electrospray ionization Q-Exactive HF-X Hybrid Quadrupole-Orbitrap mass spectrometer (ThermoFisher Scientific). Samples were separated using a 15 cm long fused silica capillary with emitter tip and frit (360 µm OD x 75 µm ID x 50 cm L, 15 µm Tip ID; MS Wil B. V., Aarle-Rixtel, The Netherlands) packed in house with ReproSil-Pur 1.9 µm C18 material (Dr. Maisch GmbH, Ammerbuch-Entringen, Germany). The peptides were separated in 0.1% FA in water (Solvent A) and 0.1% FA in ACN (Solvent B), and a column oven was attached and set to 40°C. The Whisper 20SPD method (Evosep Biosystems), corresponding to a 58-minute gradient, was used for full proteome analysis. For phosphoproteome, the Whisper 40SPD method was used (31-minute gradient).

All data was acquired in DIA mode. The methods were based on overlapping windows as described in ^62^. For full proteome analysis, the run time was set between 4 minutes and 58 minutes. The MS1 AGC target was set at 3E6 ions with a maximum injection time of 55 ms and resolution 60,000, with the scan range between 395 and 1005 m/z. DIA MS2 spectra were collected with staggered windows in 75 loops, 8 m/z isolation window, normalized collision energy (NCE) of 27, maximum injection time set to “auto” and resolution at 15,000.

For phosphoproteome acquisition, MS1 data were acquired between 4 and 31 minutes between the scan ranges of 350 and 1400 m/z, with AGC target at 3E6, 25 ms of maximum injection time and resolution 120,000. DIA MS2 data were acquired in staggered windows, with a loop count of 50, isolation window of 13.7 m/z, NCE at 27, “auto” maximum injection time and resolution of 15,000. For this method, a first fixed mass was set at 100 m/z.

### Data Processing

First, all DIA RAW files were converted to mzML with demultiplexing at 10 ppm mass error using msconvert version 3.0.21266-1f16dae (developer build)^63^. mzML files were then converted to DIA files using DIA-NN version 1.8.1^64^, which was also used for all quantitative data processing in library-free mode with 1% false discovery rate (FDR) at both peptide and protein levels.

A total of three FASTA files were used in the searches. Two corresponded to non-redundant FASTA files, namely UP000005640_9606.fasta and UP000005640_9606_additional.fasta, both downloaded in June 2023. The first file corresponds to canonical sequences from the human proteome, while the additional file contains isoforms/variants^65^. The third file used was a list of common contaminants^66^.

For full proteome searches, the precursor m/z range was set between 300 and 1800 m/z, precursor charge in the range of 1 to 4, peptide length range between 7 and 30 and a maximum of one missed cleavage was allowed. N-terminal methionine excision and cysteine carbamidomethylation were enabled. Mass accuracy was automatically set, and runs were treated as unrelated, i.e., mass accuracy and retention time scan window were determined for each file separately. “Robust LC (high-accuracy)” was used for quantification, with match between runs (MBR) enabled.

For phosphoproteome searches, the same parameters were used, with the exception that phosphorylation was enabled as a variable modification, and a maximum of 2 variable modifications were allowed.

### Bioinformatics and statistical analyses

Proteome and phosphoproteome peptide-level data were cyclic Loess normalized using NormalyzerDE. For protein-level inference, peptide identifications were subjected to protein rollup through the RRollup approach^67^ using an implementation deposited on GitHub (https://github.com/ComputationalProteomics/ProteinRollup). Briefly, the peptides, filtered at 1% FDR on DIA-NN, were grouped into protein groups with a minimum requirement of two peptides per group.

Transcriptome data for all samples was available from^20^. Gene expression Fragments per Kilobase per Million reads (FPKM) data from StringTie, adjusted to transform all data to TruSeq-like expression was used^68^. The same cyclic Loess normalization via NormalyzerDE was applied.

Samples were also accompanied by clinical data^20^, with registration at time of diagnosis from the Swedish National Quality Register for Breast Cancer (NKBC). The data included, for instance, information on receptor status, tumor grade, lymph node status, age, and follow-up data.

Immune infiltration data – estimated by histological scoring – was also available. Cellularity was determined based on hematoxylin and eosin staining of tissue microarrays created using a tissue piece adjacent to that used for nucleic acid and protein extraction. Based on the staining, scores were derived for normal epithelium, invasive and in situ tumor, lymphocyte, stroma and adipocyte content ^18^.

All statistical analyses were performed in RStudio, using R version 4.3.3. A Nextflow workflow was implemented to run all steps of the analysis and generate all figures. A graphical representation of such workflow can be seen in **Figure 6**. All source code is available on GitHub: https://github.com/ComputationalProteomics/BreastCancerMultiomics. A Docker container with all dependencies installed is available.

**Figure 6.**
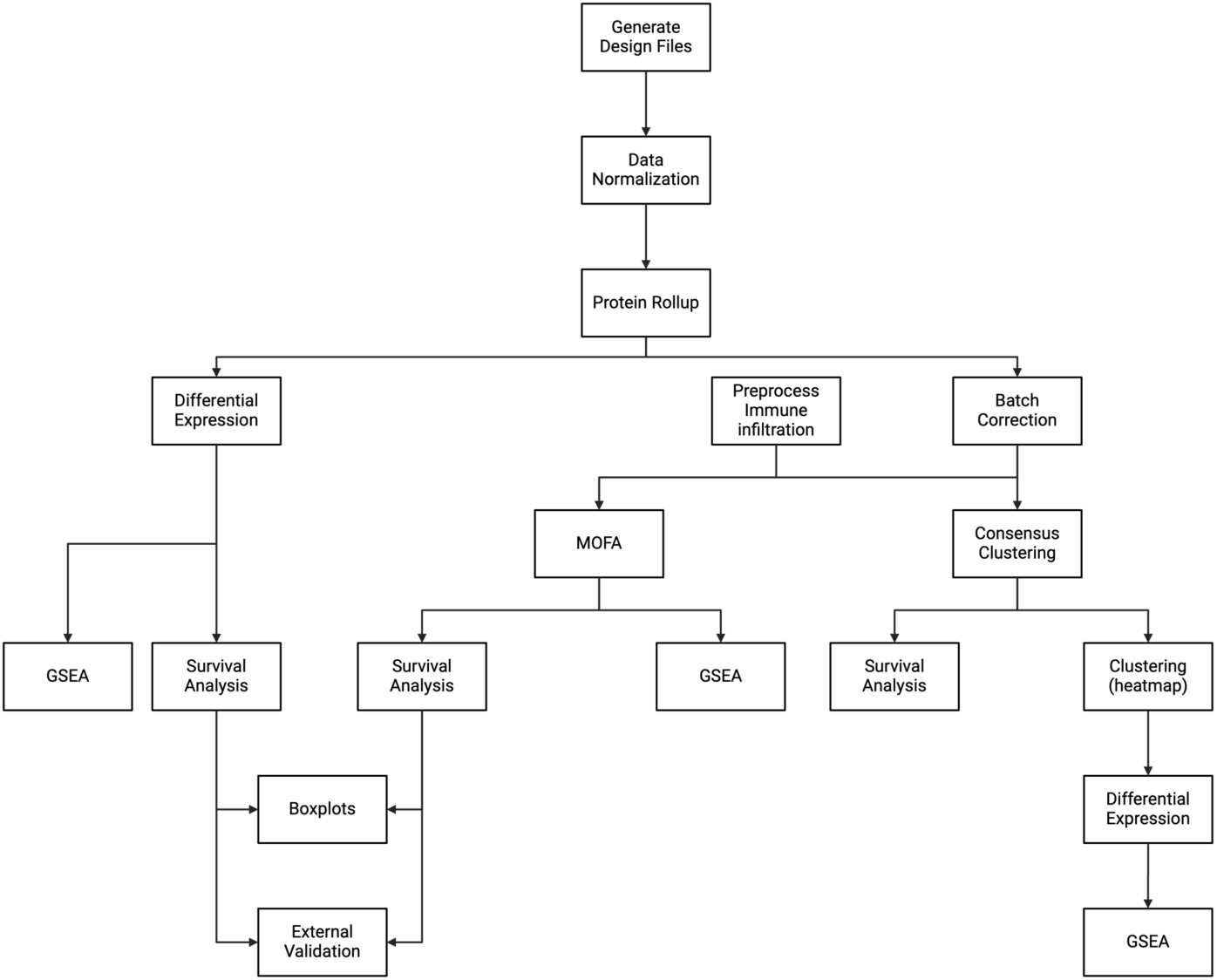
Data analysis workflow. Created with BioRender.com.

#### Differential Expression Analysis

Differential expression analysis between different conditions was performed at the protein level for full proteome data, peptide level for phosphoproteome data and transcript level for transcriptome data. It was conducted via LIMMA statistics implemented in NormalyzerDE.

#### Gene Set Enrichment Analysis

Gene set enrichment analysis (GSEA) was performed via the GSEA() function implemented in ClusterProfiler^69^ using a ranked list of features based on differential abundance between different conditions and omics. The list was ranked based on the -log10 of the p-value with the added sign from the log2 fold change values. All parameters were kept at their default settings, apart from the p-value adjustment method (pAdjustMethod = ‘fdr’) and q-value cutoff (pvalueCutoff=0.25). The ranked list was searched against the Hallmark gene set from the Molecular Signatures Database (MSigDB)^70–72^. For PTM Signature Enrichment Analysis (PTM-SEA)^34^ of the phosphoproteome data, the quantitative phosphopeptides data were first aggregated per phosphosite, with mean intensities of phosphopeptides covering the same phosphosite. PTM-SEA was then run using flanking sequence phosphosite annotation with ranking based on signed log10 p-values of group comparisons (as for the GSEA) using the ssGSEA2.0 implementation downloaded from https://github.com/broadinstitute/ssGSEA2.0 with the complete list of ptmsigdb 2.0 human signatures. For illustrations the PTM signatures passing FDR<0.01 in any comparison were plotted.

#### Unsupervised Consensus Clustering

Consensus clustering was performed with the ConsensusClusterPlus R package^73^. Incomplete features were removed from, and the 25% most variable features at the protein and transcript levels (based on median absolute deviation) were used. For phosphoproteome data, the 50% most variable peptides were used. A total of 1000 repeats of the Partitioning Around Medoids (PAM) algorithm was run with Pearson correlation as the distance metrics and a maximum number of clusters of 12 (maxK=12).

The optimal number of clusters for each omics was decided based on inspection of correlation matrices, consensus Cumulative Distribution Function (CDF) plots and delta area plots.

For visualization of the clustering results, the ComplexHeatmap R package was used^74^.

After removal of incomplete features, the top 15%, 25% and 75% most variable features (based on MAD) were kept for transcriptome, proteome and phosphoproteome data, respectively. All feature values were then converted to Z-scores prior to clustering.

Features were clustered in 5 distinct clusters using 1000 repetitions of K-means clustering when generating the heatmap. For each block, functional annotation was added. The annotations were generating by performing g:Profiler enrichment analysis in the gprofiler2 R package^75^. The following parameters were used: organism = ‘hsapiens’, multi_query = FALSE, significant = TRUE, ordered_query = FALSE, exclude_iea = TRUE, measure_underrepresentation = FALSE, evcodes = TRUE, user_threshold = 0.1, correction_method = ‘gSCS’, custom_bg = NULL, numeric_ns = “”, domain_scope = ‘annotated’, sources = NULL. The results were then filtered to keep sources in GO:BP, GO:MF or REACTOME, term size below 150 for transcriptome and proteome data and below 600 for phosphopeptide data. Finally, the 5 terms with lowest p-value were selected and added to the heatmaps.

To give an overview of the cluster membership of samples across the different omics layers, an alluvial plot was generated using the ggalluvial R package ^76^.

#### Estimation of Immune Infiltration

Immune cell infiltration was estimated using the CIBERSORTx ^26^ and EPIC ^27^ algorithms. For CIBERSORTx analysis, the CIBERSORTx web tool was employed as the Docker container does not support using absolute mode. EPIC was run via the immunedeconv R package ^77^. Prior to running CIBERSORTx or EPIC, the protein data was subjected to several preprocessing steps. Briefly, HGNC gene symbols were updated to match current nomenclature, protein groups and gene groups were condensed into the first protein or gene in each group, missing values were imputed with the minimum value found in the dataset, duplicate occurrences of genes were removed (preferring the gene with overall highest intensities) and scaling was performed in a TPM-like manner. For EPIC, the built-in transcriptome-based signature based on tumor-infiltrating lymphocytes was used. For CIBERSORTx, either the LM22 matrix derived from microarray data, or a custom matrix derived from proteome data was used.

#### Multi Omics Factor Analysis

Multi omics Factor Analysis (MOFA) was performed using the MOFA2 R package^25^. For this integrative omics analysis, proteome, phosphopeptide and transcriptome data were used. First, the data were filtered to only keep features with a minimum of 70% data completeness, defined by a cutoff of maximum 30% for incomplete observations, as well as keeping the 25% most variable features based on median absolute deviation. Immune infiltration data based on two deconvolution algorithms, EPIC and CIBERSORTx, were also included as separate omics layers. Each omics layer, referred as views in MOFA documentation, was scaled prior to model training.

All models were generated based on 20 factors, with spike and slab sparsity and ARD sparsity enabled (model_opts$spikeslab_weights = TRUE, model_opts$ard_weights = TRUE). Default training options were used, with convergence mode set to “slow”. For each model generated, 10 different models were trained with different seeds and the best one was selected using the select_best() function from the MOFA2 package, which selects the best model based on the best ELBO value.

#### Survival Analysis

Survival analysis was performed using the censored extension of the tidymodels meta-package ^78,79^. The proportional_hazards() model was used with the survival package as the engine and “censored regression” as the mode.

Two different approaches were used for performing survival analyses in the identification of relevant features, the first originating from features based on the differential expression analyses across different omics, and the second based on factors in the MOFA models which significantly correlated to the outcome of interest. For the survival endpoints, overall survival (OS), recurrence-free interval (RFi), and distant recurrence-free interval (DRFi) were used ^20,80,81^. Features which were deemed significant in univariable model, and which did not violate the proportional hazards assumption – defined by non-significant (p-value>0.05) results based on the cox.zph function – were tested in a multivariable model, compensating for age at diagnosis, tumor size, grade, lymph node status and treatment. Forest plots were generated using the forest_model() function from the forestmodel R package.

For the first approach, differentially abundant features from all three omics layers were filtered based on adjusted p-value (adj. p-value < 0.1) followed by removal of features with incomplete observations.

For survival analysis based on MOFA models, factors correlated with the clinical outcome of interest, i.e., lymph node involvement or presence of distant metastasis, were selected (adjusted p-value < 0.05). Features with an absolute weight greater than 0.5 in any of these factors were chosen.

Cox proportional hazards was also used to investigate whether the clusters defined by consensus clustering analysis had differential survival outcomes. Forest plots based on univariable analysis were generated using the ggplot2 package.

#### External Validation

Candidates deemed significant at both univariable, and multivariable survival analysis were further validated using the Human Protein Atlas ^29,30^ and the Kaplan-Meier Plotter^31,32^.

Survival analysis reported on the Human Protein Atlas is based on the FPKM value of each gene. Patient group prognosis is determined by Kaplan-Meier plots and log-rank tests. Both median and maximally separated values are reported. Worth mentioning, genes with log-rank p-values under 0.001 are considered prognostic, with the survival of the high expression group determining whether it is favorable or unfavorable. Moreover, lowly expressed genes, defined by median expression below FPKM 1 are classified as unprognostic regardless of the survival analysis results.

Kaplan-Meier Plotter ^82^ survival analysis was performed at the gene level, using median expression for cutoff

An additional external validation was conducted based on published transcriptome data from SCAN-B samples^20,68^. Significant features from either univariable or multivariable analysis were selected and mapped to gene identifiers found in the transcriptome data. Gene expression data from ER-positive/HER2-negative samples not included in the present study and collected latest in 2016 underwent the same Loess normalization described previously, followed by analysis via univariable and multivariable Cox proportional hazards models as described previously.

## Funding

The study was supported by the Erling Persson Foundation and the European Union’s Horizon 2020 research and innovation programme under the Marie Skłodowska-Curie grant agreement No. 754299 (EU-H2020-MSCA-COFUND-754299-CanFaster).

## Acknowledgements

The authors would like to acknowledge all patients and healthcare providers participating in the SCAN-B study, personnel at the central SCAN-B laboratory at the Division of Oncology, Department of Clinical Sciences Lund, Lund University, the Swedish national breast cancer quality registry (NKBC), Regional Cancer Center South, RBC Syd, and the South Sweden Breast Cancer Group (SSBCG) and the Kamprad Foundation. Pär-Ola Bendahl is thanked for discussions regarding sample selection.

## Conflicts of Interest

The authors declare no conflict of interest.

## Data Availability

The generated mass spectrometry proteomics and phosphoproteomics data have been deposited on the ProteomeXchange Consortium via the PRIDE partner repository with the dataset identifier PXD059920.

## Code Availability

All source code is available on GitHub: https://github.com/ComputationalProteomics/BreastCancerMultiomics. A Docker container with all dependencies installed is available.

## Supplementary Figures

**Supplementary Figure S1.**
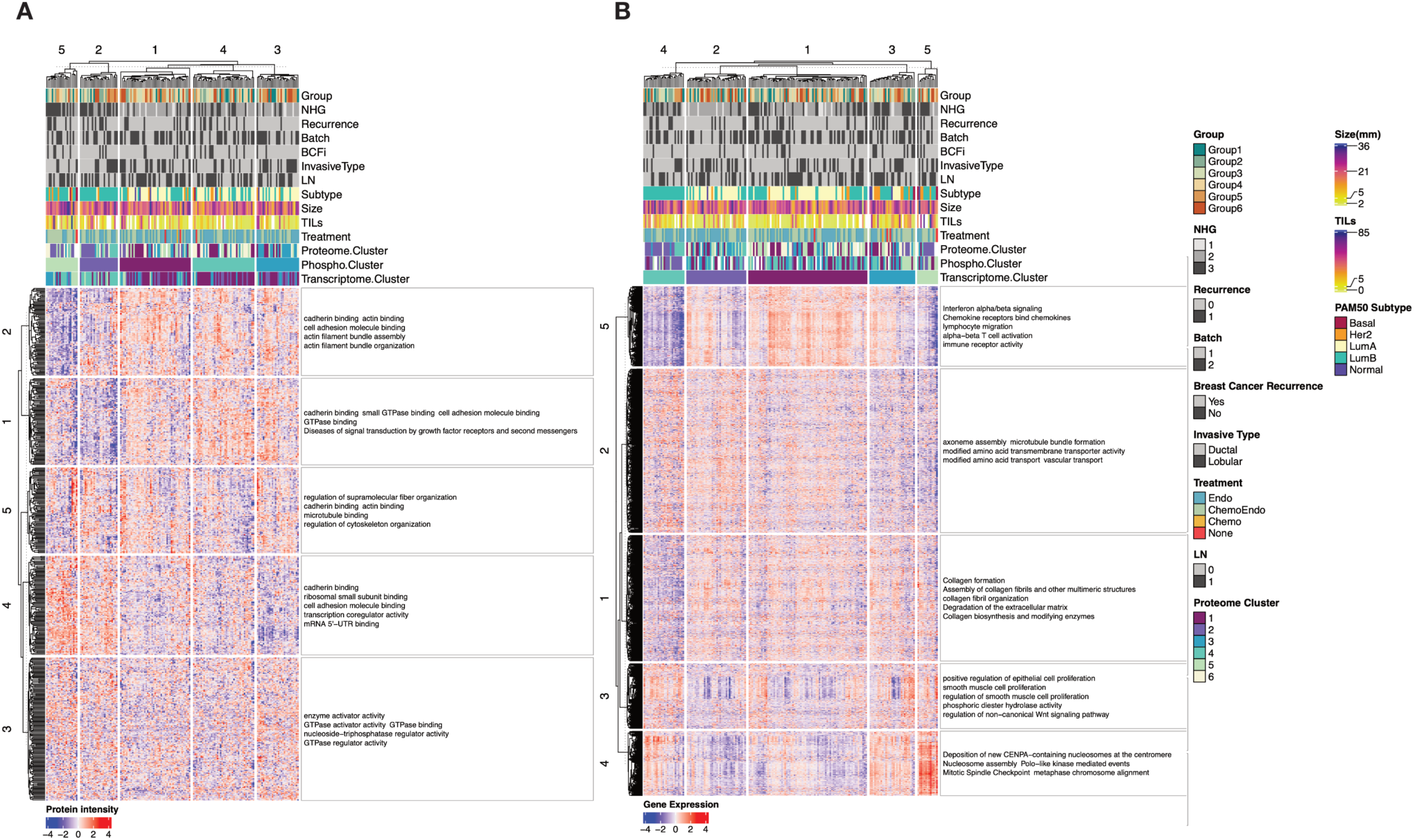
Consensus clustering at the phosphoproteome **(A)** and transcriptome **(B)** level. The features were further clustered in 5 distinct clusters based on K-means and the terms represent enrichment based on G:profiler.

**Supplementary Figure S2.**
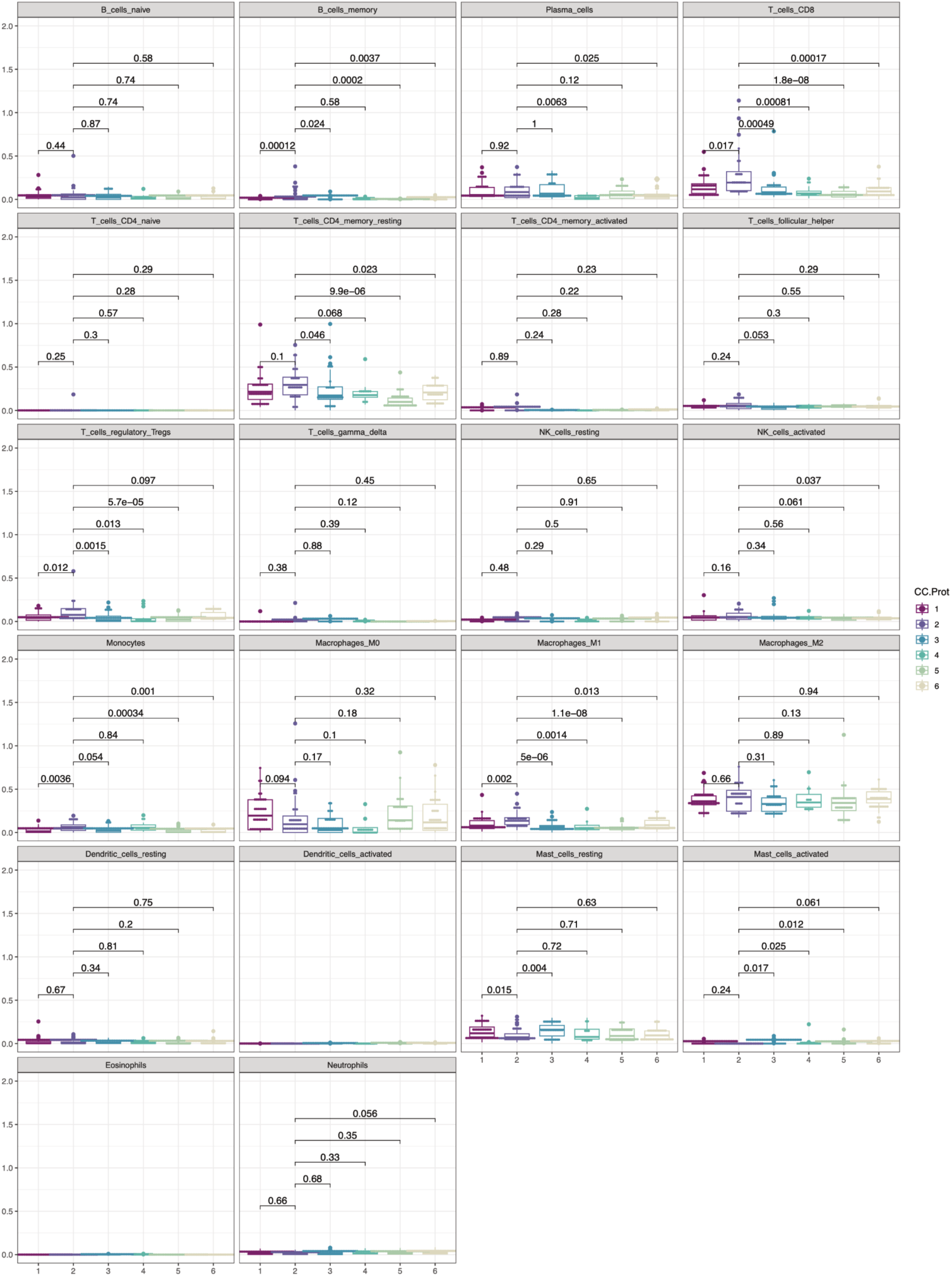
Comparison of immune infiltration estimates based on CibersortX using absolute mode and LM22 signature matrix. P-values are based on Wilcoxon test.

## Supplementary Tables

**Supplementary Table 1: Differentially abundant proteins in NST vs ILC at FDR<0.05 and absolute log2 Fold Change > 1.**

**Supplementary Table 2: Summary of univariable and multivariable Cox analysis based on RFS and OS for features identified in the MOFA model and associated with lymph node involvement.**

**Supplementary Table 3: Summary of univariable and multivariable Cox analysis based on DRFS and OS for features identified in differential expression analysis and associated with distant metastasis.**

**Supplementary Table 4: Summary of univariable and multivariable Cox analysis based on DRFS and OS for features selected in MOFA model and associated with distant metastasis.**

## Notes

### Competing Interest Statement

The authors have declared no competing interest.

### Author Declarations

Included patients were enrolled in the Sweden Cancerome Analysis Network - Breast (SCAN-B) study (ClinicalTrials.gov ID NCT02306096) (PMID:25722745,29341157) approved by the Regional Ethical Review Board in Lund, Sweden (registration numbers 2009/658, 2010/383, 2012/58, 2013/459, 2014/521, 2015/277, 2016/541, 2016/742, 2016/944, 2018/267 and the Swedish Ethical Review Authority (registration numbers 2019-01252, 2024-02040-02).

